# Evaluation of gonadal function in transfusion-dependent β-thalassemia patients of Bangladesh

**DOI:** 10.1101/2022.01.22.22269688

**Authors:** Romana Chowdhury, Mohammad Mehedi Hasan, Mushfiq Newaz Ahmed, Mohammad Azmain Iktidar, Md. Mazharul Hoque Tapan, Sheikh Saiful Islam Shaheen, Atiar Rahman, Ayesha Khatun

## Abstract

**Background:** Hypogonadism is one of the most frequent complications in transfusion-dependent thalassemia patients, and early recognition and treatment is the core element to restore impaired gonadal function. Despite the high burden of disease, relevant studies are scarcely addressing the gonadal function of such patients. The pattern of gonadal function in transfusion-dependent thalassemia patients must be picturized before planning a generalized management plan; therefore, this study was conducted.

**Methods:** This cross-sectional study was conducted at the Department of Transfusion Medicine of Bangabandhu Sheikh Mujib Medical University. According to inclusion and exclusion criteria, a total of 94 patients were enrolled in this study. A detailed history and thorough clinical examination were carried out in each patient and recorded using a pretested structured questionnaire. In addition, laboratory assessment of LH, FSH, testosterone and estradiol in serum were also done. Data were analyzed using STATA (v.16).

**Results:** The mean age of the patients with transfusion-dependent thalassemia was 18.81±4.65 (SD), with 53.3% of the patients being male. The most common symptoms of gonadal dysfunction among males were loss of body hair (6%) followed by fatigue (4%), and among females were slow or absent breast growth, hot flashes and amenorrhea (6.82% each).

**Conclusion:** The overall prevalence of hypogonadism was 35.11%, 18.1% being normogonadotropic, 11.7% being hypogonadotropic, and 5.3% being hypergonadotropic. Therefore, gonadal hormone analysis of transfusion-dependent thalassemia patients can be considered a screening tool for assessing gonadal function and early detection and prevention of hypogonadism.

## Introduction

Thalassemia is one of the most common genetic hemoglobinopathies that can result in severe anemia [1,2]. It is highly prevalent in Southeast Asia, the Indian subcontinent, Mediterranean, and Middle Eastern countries, collectively known as the ‘World Thalassemia Belt’. In Bangladesh, 6–12% of the population are carriers of a gene causing thalassemia [3]. Thalassemia is characterized by the partial or complete deficiency in the synthesis of α or β-globin chains that compose the major adult hemoglobin (α_2_β_2_). Patients with an absolute deficiency of the α or β-globin chains require lifelong blood transfusion and are denoted as transfusion-dependent thalassemia group [4]. With regular transfusion, the life expectancy and survival rate of thalassemia patients dramatically improved from 1^st^ decade of life to 5^th^ decade [5]. However, it leads to iron overload, which is accumulated within different tissues, including endocrine glands resulting in functional imbalance, among which gonadal dysfunction is the most common [6,7]. Besides iron overload, factors such as ferritin level, genotype, transfusion frequency, starting age and iron chelation efficiency also play a significant role [8]. Hypogonadotropic hypogonadism or secondary hypogonadism resulting from iron deposition in the pituitary gonadotrope is more commonly found, whereas gonadal iron deposition in ovaries or testes occurs less frequently [9].

In female patients, gonadal hormones such as luteinizing hormone (LH), estradiol, follicle-stimulating hormone (FSH), anti-mullerian hormone (AMH), prolactin are lower in transfusion-dependent thalassemia patients [7,10]. Nearly half of these patients show a low to undetectable LH/FSH ratio [11]. As a result, amenorrhea, anovulation, and infertility are commonly found in adults. In younger females, the low gonadal function manifests as delayed puberty, delayed menarche or primary amenorrhea, and short stature [12]. In male patients, testicular functions are seen to be reduced, evidenced by high anti-mullerian hormone and low testosterone levels [13]. Lower sperm concentrations, a lower percentage of sperm with normal morphology, sperm DNA damage, reduced testis volume are also seen in transfusion-dependent thalassemia patients [14]. The adult male’s clinical features are loss of facial and body hair, decreased muscle mass, and appearance of fine facial wrinkles, gynecomastia, diminished libido, fatigue, ejaculatory dysfunction [12,15].

Early assessment of gonadal dysfunction and its prompt treatment by chelation therapy may reduce its incidence, improve the quality of life in already burdened thalassemic patients and prevent further complications [8]. Despite the high prevalence of thalassemia in our context, baseline information of gonadal function is not well addressed. Therefore, this study aimed to find the status of gonadal function in transfusion-dependent thalassemia patients.

## Materials and Methods

### Study Design, Site & Duration

This cross-sectional study was conducted from June 2020 to June 2021. Department of Transfusion Medicine, Bangabandhu Sheikh Mujib Medical University (BSMMU), Dhaka, Bangladesh, was selected as the study site as it is located in the capital and receives patients from all over the country.

### Study Participants

β-thalassemia patients diagnosed by hemoglobin electrophoresis, who were dependent on transfusion and providing consent, were included in the study. β-thalassemia patients with other chronic illnesses that can cause gonadal dysfunction (e.g., connective tissue disease), under hormonal replacement therapy or taking OCP, with congenital gonadal malformations, taking a regular antipsychotic, antidepressant and other drugs that hamper gonadal function were excluded from the study. A total of 400 transfusion-dependent β-thalassemia patients were sampled by judgment type of non-probability sampling. Among which 277 were excluded for meeting one or more of the exclusion criteria, and 29 were excluded as they/their guardian didn’t provide consent to enter the study, leaving a final sample size of 94.

### Data Collection

Data were collected by face-to-face interviews of patients/guardians using a pretested structured questionnaire. Background information, previous medical records, physical findings, and laboratory reports were also assessed. The gonadal function of the patient was evaluated by history, physical examination and laboratory tests, including serum luteinizing hormone (LH), serum follicle-stimulating hormone (FSH), serum testosterone for males and serum estradiol for females. Blood samples (3-5 ml) were collected from the patients put into labelled test tubes, and left for clotting with all available aseptic precautions. The serum was then separated by centrifugation at 6000 rpm after the blood had clotted. Assessment of hormone levels was done in the Department of Biochemistry and Microbiology, BSMMU, by VEG-4000 Fully Automated ELISA Processor (manufactured by ESSE 3 medical equipment, Italy).

### Statistical Analysis

After data collection, data were checked for errors and analyzed using STATA (version 16). Continuous variables were presented as mean and standard deviation, and categorical variables were presented as frequency and relative percentage. In addition, Pearson’s chi-square test was performed to explore a bivariate relationship. A *P-value* of <0.05 is considered statistically significant, and all the reporting is done according to the STROBE guidelines[16].

### Ethics

This study was approved by the Institutional Review Board of BSMMU. The 1964 Declaration of Helsinki and later modifications and comparable ethical standards were followed wherever feasible. We promised respondents that we would never reveal any portion of the interview to an unauthorized individual.

## Results

The mean age of the participants was 18.81±4.65 years, with the maximum being between 13 to 18 years of age (60.6%). There was a male (53.2%) preponderance with a male to female ratio of 1.14:1. Most of the patients were from rural area (57.4%) [Table 1].

**Table 1:** Background information of study participants (N=94)

| Characteristics | Entire Study Cohort (N=94) |
| --- | --- |
| **Age (in years), mean ± SD** | 18.81±4.65 |
| 13 to 18 | 57 (60.64) |
| 19 to 24 | 24 (25.53) |
| 25 to 30 | 13 (13.83) |
| **Gender** |  |
| Male | 50 (53.19) |
| Female | 44 (46.81) |
| **Residence** |  |
| Rural | 27 (54) |
| Urban | 23 (46) |

Values are expressed as n(%) unless otherwise mentioned
Abbreviations: SD, standard deviation;

The most common symptoms of gonadal dysfunction among males were loss of body hair (6%) followed by fatigue (4%), and among females were slow or absent breast growth, hot flashes and amenorrhea (6.82% each) [Table 2].

**Table 2:**
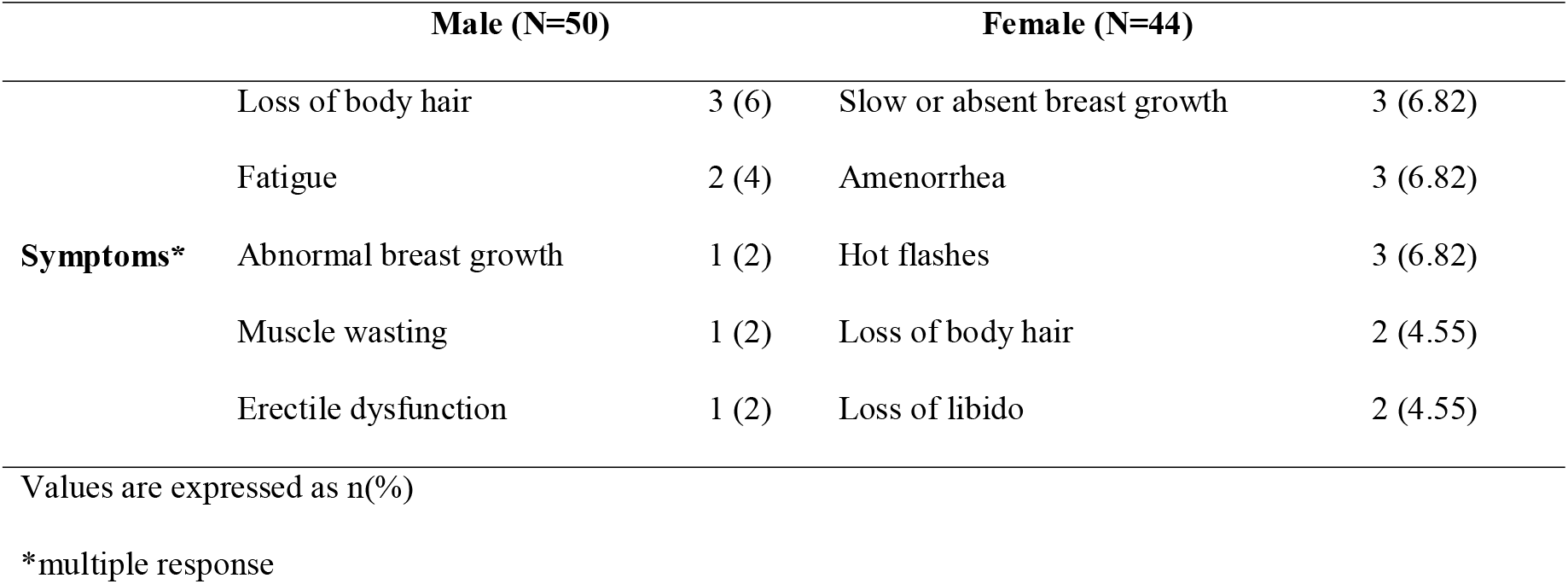
Symptoms of gonadal dysfunction among study participants (N=94)

Amongst 94 patients, the mean serum FSH of all patients was 4.52±2.83 IU/L, wherein the majority (89.36%) had a normal level of FSH, and only 8.51% of patients had below the normal level of FSH. Among the male, 4% had above the normal level of FSH, and none had below the normal level of FSH. Among females, 18.18% had a below normal level of FSH, and none had above the normal level of FSH [Table 3]. The mean serum LH of all patients was 3.93±2.82 IU/L wherein the majority (76.60%) had a normal level of FSH, and 11.70% had below and above the normal level of LH each. Among the male, 22% had an above normal level of LH, and 10% had a below normal level of LH. Among females, 13.64% had below normal LH, and none had above normal level of LH. Among all the male patients, mean serum testosterone was 390.29±287.93 ng/dL wherein the majority (64%) had a normal level of testosterone and the rest 36% patients had below normal testosterone [Table 3]. The mean serum estradiol of female patients was 85.05±153.47 pg/mL, whereas the maximum (52.27%) had a normal estradiol level. Among rest, 13.64% had above the normal level of estradiol, and 34.09 % of patients had below the normal level of estradiol.

**Table 3:** Laboratory findings for gonadal profile in study participants (N=94)

| **Hormones** | **Male**<br>**N=50** | **Female**<br>**N=44** | **Total**<br>**N=94** |
| --- | --- | --- | --- |
| **S. FSH (IU/L), mean±SD** | 4.58±3.34 | 4.44±2.16 | 4.52±2.83 |
| Below normal | 0 (0) | 8 (18.18) | 8 (8.51) |
| Within normal | 48 (96) | 36 (81.82) | 84 (89.36) |
| Above normal | 2 (4) | 0 (0) | 2 (2.13) |
| **S. LH (IU/L), mean±SD** | 4.19±3.4 | 3.63±1.97 | 3.93±2.82 |
| Below normal | 5 (10) | 6 (13.64) | 11 (11.70) |
| Within normal | 34 (68) | 38 (86.36) | 72 (76.60) |
| Above normal | 11 (22) | 0 (0) | 11 (11.70) |
| **S. Testosterone (ng/dL), mean±SD** | 390.29±287.93 | - | - |
| Below normal | 18 (36) | - | - |
| Within normal | 32 (64) | - | - |
| Above normal | 0 (0) | - | - |
| **S. Estradiol (pg/mL), mean±SD** | - | 85.05±153.47 | - |
| Below normal | - | 15 (34.09) | - |
| Within normal | - | 23 (52.27) | - |
| Above normal | - | 6 (13.64) | - |

Values are expressed as n(%) unless otherwise mentioned
Abbreviations: S., serum; SD, standard deviation; FSH, follicle-stimulating hormone; LH, luteinizing hormone.

Hypogonadism was found in 33 patients (35.11%), with 36% of all male and 34% of all female patients suffering from hypogonadism. Normogonadotropic hypogonadism was the most prevalent type 17 (18%), followed by hypogonadotropic hypogonadism 11 (12%) and hypergonadotropic hypogonadism 5 (5%). The majority of the male patients with hypogonadism had normogonadotropic type 10 (20%); in contrast, majority of the female patients with hypogonadism had hypogonadotropic type 8 (18%). There was a significant difference in the distribution of types of hypogonadism between males and females (*P*=0.023) [Fig. 2].

**Fig. 1:**
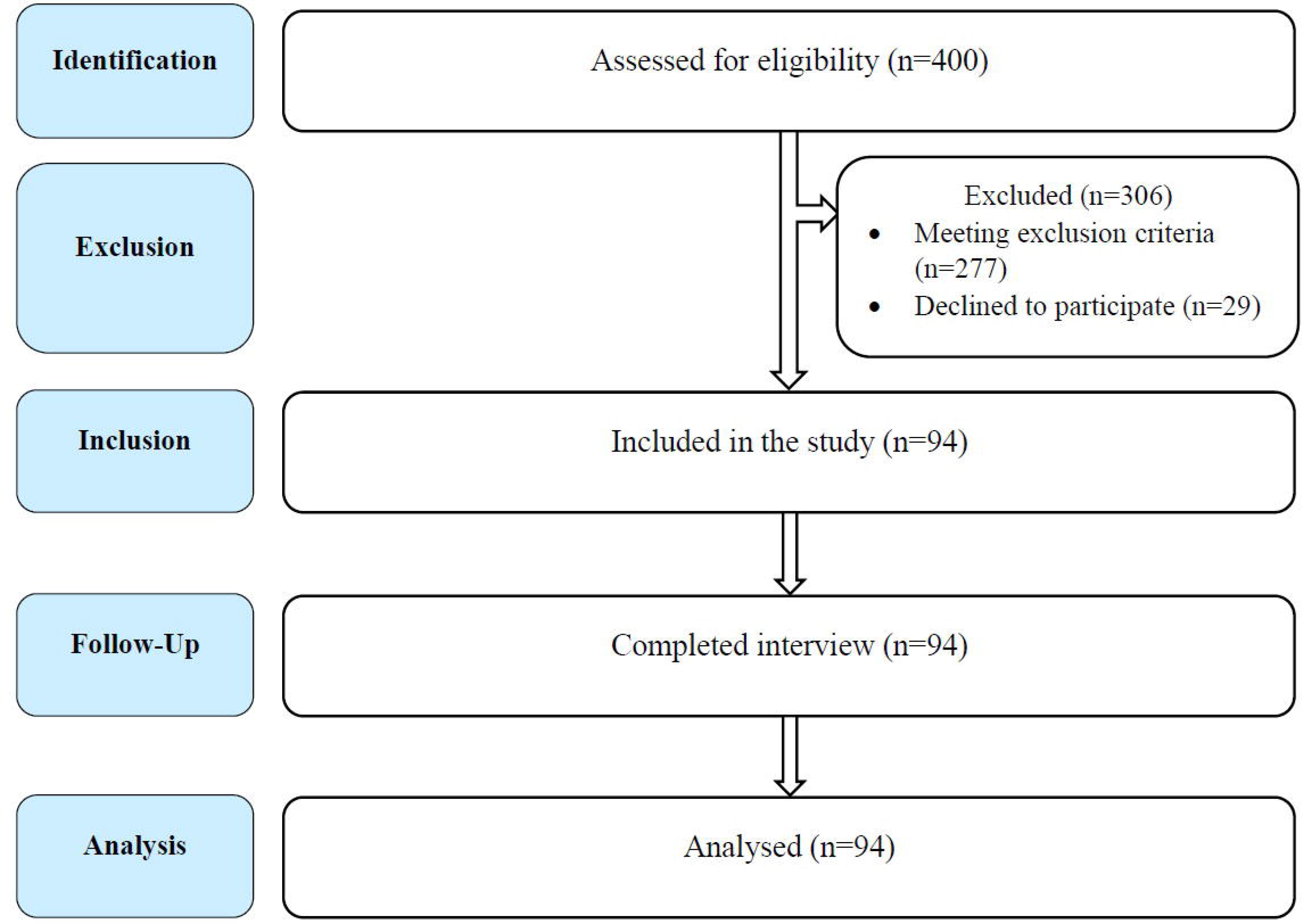
STrengthening the Reporting of OBservational studies in Epidemiology (STROBE) flow chart of study participants.

**Fig. 2:**
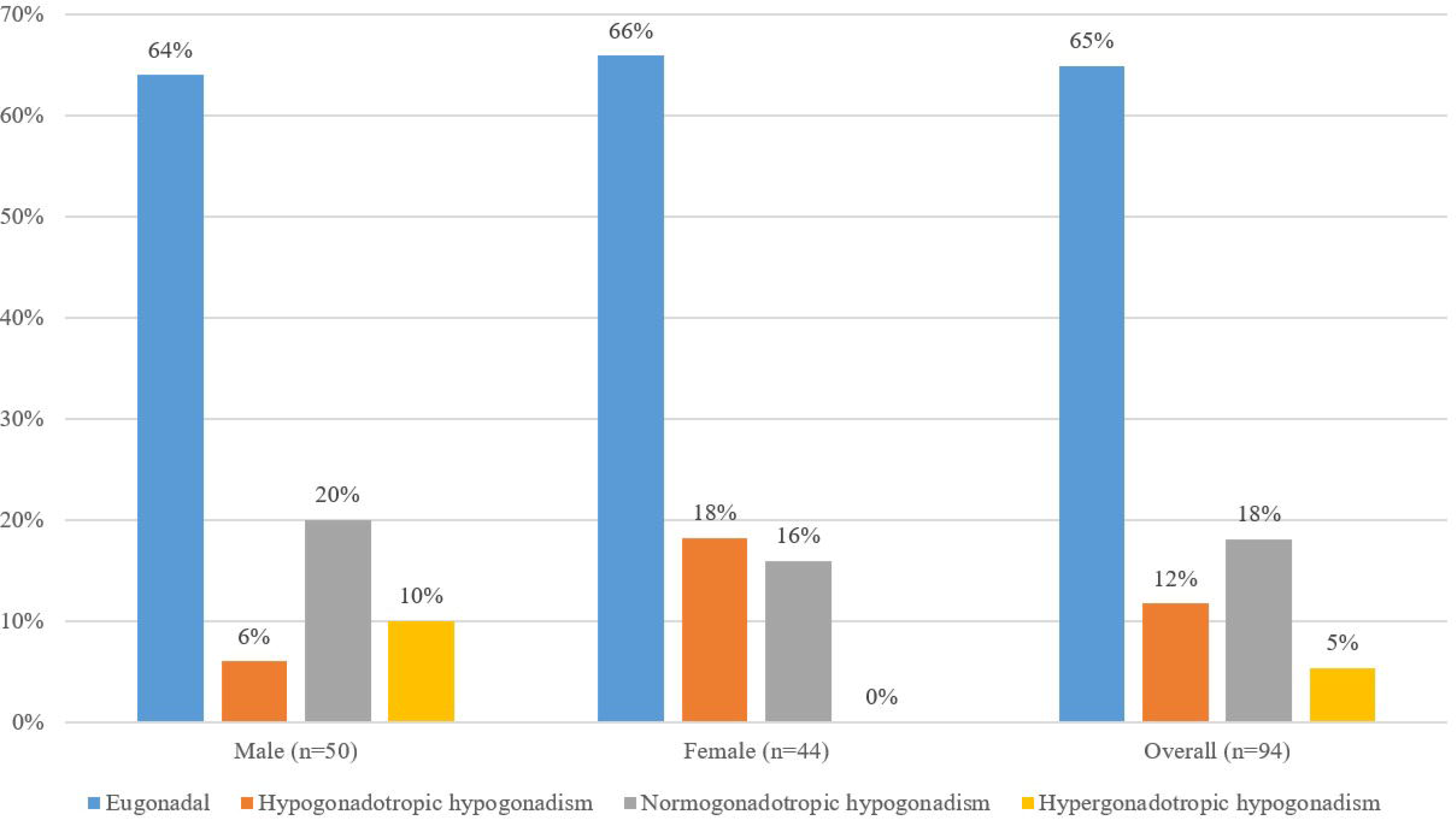
Status of gonadal function among the study participants. Pearson’s chi-square test revealed significant difference in distribution of types of hypogonadism between male and female (p=0.023)

## Discussion

Transfusion-dependent beta-thalassemia, also known as beta-thalassemia major (β-TM), most often require regular red blood cell transfusions, which start within the first year of life (Chern et al. 2003; Chen et al. 2018). The predilection for iron deposition in the pituitary gland and hypothalamus among these patients frequently causes a lack of sexual maturation and loss of gonadal function. However, controversy still exists with respect to the causes of gonadal dysfunction due to primary iron deposits in the gonads or secondary to a hypogonadotropic state in patients with beta-thalassemia major.

In our study, the overall prevalence of hypogonadism was 35.11%, 18.1% being normogonadotropic, 11.7% being hypogonadotropic, and 5.3% being hypergonadotropic. In a study among 21 male patients with β-TM, the prevalence of hypogonadotropic hypogonadism was 33.3%, which is consistent with our study [18]. Also, a cohort study in Taiwan among 454 β-TM major patients showed a lower prevalence of hypogonadism (23.1%) [19]. Another study carried out by Daraghmeh et al., in the thalassemia ward at AL-Wattani hospital in Nablus among 75 β-TM patients found that hypogonadism was prevalent among 46.7% of the tested patients, including both primary and secondary hypogonadism [20]. However, the prevalence of hypogonadism in this study population was lower than some published reports [7,10,21–23] all of which have reported hypogonadism as the most frequent endocrinopathy among β-TM patients from different countries. The availability of iron chelation therapy can explain the lower prevalence of hypogonadism in our study compared to other studies. A study on 382 β-TM patients treated with desferrioxamine at the Thalassemia Centre in Dubai showed a significantly lower prevalence of hypogonadism of only 25% [23]. Besides, chelation therapy with desferrioxamine before puberty has helped patients attain normal sexual maturation in some studies. In a study of 40 patients with β-TM, 90% of 19 patients who began treatment with desferrioxamine before the age of 10 years had normal sexual development compared with only 38% of those treated after the age of 10 years [24]. However, results of a prospective study in patients with thalassemia and secondary amenorrhea suggest that the damage to the hypothalamus is progressive, despite regular transfusion and chelation therapy [25]. These results suggest that the development of hypogonadotropic hypogonadism might be caused by early and progressive damage due to iron loading. We believe that these could be attributed to the possible progressive toxic effect of iron-induced free radicals and/or to some other undefined risk factors involved in the development of hypogonadotropic hypogonadism [17].

There was no statistically significant relationship between gender and hypogonadism in our study. This result was similar to the studies carried out in Iran and Palestine [22,26]. However, this result was different from the results of a study by Dumaidi et al., which stated that a higher percentage of hypogonadism in males indicates that males are more prone to hypogonadism than females [10].

In the current study, serum testosterone, estradiol, LH and FSH were lower among 36%, 34.09%, 11.70% and 8.51% patients, respectively. The most common symptom of gonadal dysfunction among males was the loss of axillary and pubic hair (6%), and among females was slow or absent breast growth, hot flashes and amenorrhea (6.82% each). Regarding the menstrual status, adult females presented hypogonadism in the form of total absence of spontaneous menarche (primary amenorrhea): this applied to 29% of female participants with β-TM, as confirmed by the low level of estradiol;. In contrast, the other form is lack or irregular menses (secondary amenorrhea) was found in 42% of female participants with β-TM. In males, hypogonadism was in the form of sexual infantilism: this applied to 72% of male participants with β-TM, which agrees with contemporary studies [27,28]. Several other studies also revealed that, in the thalassemic group, the baseline and peak levels, after the GnRH test, of luteinizing hormone (LH), follicle-stimulating hormone (FSH), and estradiol were significantly lower than those in the control group [29][30]. However, the discrepancy in gonadotropin hormone level and gonadal dysfunction symptoms among males and females compared to other studies might be explained by ethnic factors, different availability of therapeutic agents and economic status [7].

In our study, approximately one-third of the transfusion-dependent β-thalassemia patients were found to be suffering from different hypogonadism. Normogonadotropic hypogonadism was the most prevalent, followed by hypogonadotropic and hypergonadotropic. Therefore, gonadal hormone analysis of transfusion-dependent thalassemia patients can be considered a screening tool for assessing gonadal function and early detection and prevention of hypogonadism. However, further prospective studies with a larger sample size might be necessary to provide better recommendations for patients with transfusion-dependent β-thalassemia.

## Data Availability

All data produced in the present work are contained in the manuscript

## Acknowledgment

We want to thank all the patients who participated in this study.

## References

[1] Ansari S, Kiumarsi A, Azarkeivan A, Allameh MM, Amir kashani D, Razaghi Azar M. Fertility assessment in thalassemic men. Thalass Reports 2017;7:21–4. https://doi.org/10.4081/thal.2017.6362.

[2] Mensi L, Borroni R, Reschini M, Cassinerio E, Vegetti W, Baldini M, et al. Oocyte quality in women with thalassaemia major: insights from IVF cycles. Eur J Obstet Gynecol Reprod Biol X 2019;3. https://doi.org/10.1016/j.eurox.2019.100048.

[3] Hossain MS, Hasan MM, Raheem E, Islam MS, Al Mosabbir A, Petrou M, et al. Lack of knowledge and misperceptions about thalassaemia among college students in Bangladesh: a cross-sectional baseline study. Orphanet J Rare Dis 2020;15:54. https://doi.org/10.1186/s13023-020-1323-y.

[4] Taher AT, Weatherall DJ, Cappellini MD. Thalassaemia. Lancet 2018;391:155–67. https://doi.org/10.1016/S0140-6736(17)31822-6.

[5] Cao A, Moi P, Galanello R. Recent Advances in β-Thalassemias. Pediatr Reports 2011, Vol 3, Pages 65-78 2011;3:65–78. https://doi.org/10.4081/PR.2011.E17.

[6] Al-Zuhairy Tnahs, Al-Ali Zajr. Evaluation of reproductive hormones in patients with β-thalassemia major in misan province, iraq. Medico-Legal Updat 2020;20:274–9.

[7] Albu AI, Albu D. Hypogonadism in Female Patients with Beta Thalassemia Major. Thalass Other Hemolytic Anemias 2018. https://doi.org/10.5772/intechopen.73862.

[8] De Sanctis V, Soliman AT, Elsedfy H, Di Maio S, Canatan D, Soliman N, et al. Gonadal dysfunction in adult male patients with thalassemia major: an update for clinicians caring for thalassemia. Expert Rev Hematol 2017;10:1095–106. https://doi.org/10.1080/17474086.2017.1398080.

[9] Srisukh S, Ongphiphadhanakul B, Bunnag P. Hypogonadism in thalassemia major patients. J Clin Transl Endocrinol 2016;5:42–5. https://doi.org/10.1016/j.jcte.2016.08.001.

[10] Dumaidi K, Al-Jawabreh A, Al-Assi S, Karmi B. Assessment of gonadal and thyroid function for adult transfusion-dependent-β-thalassemic patients in Palestine. Jordan Med J 2015;49:17–26. https://doi.org/10.12816/0025095.

[11] Singer ST, Sweeters N, Vega O, Higa A, Vichinsky E, Cedars M. Fertility potential in thalassemia major women: current findings and future diagnostic tools. Ann N Y Acad Sci 2010;1202:226–30. https://doi.org/10.1111/j.1749-6632.2010.05583.x.

[12] De Sanctis V, Elsedfy H. Clinical and Biochemical Data of Adult Thalassemia Major patients (TM) with Multiple Endocrine Complications (MEC) versus TM Patients with Normal Endocrine Functions: A long-term Retrospective Study (40 years) in a Tertiary Care Center in Italy. Mediterr J Hematol Infect Dis 2016;8:e2016022. https://doi.org/10.4084/mjhid.2016.022.

[13] Siripunthana S, Sahakitrungruang T, Wacharasindhu S, Sosothikul D, Supornsilchai V. Testicular function in patients with regular blood transfusion for thalassemia major. Asian Biomed 2015;9:185–91. https://doi.org/10.5372/1905-7415.0902.385.

[14] Chen M-J, Peng SS-F, Lu M-Y, Yang Y-L, Jou S-T, Chang H-H, et al. Effect of iron overload on impaired fertility in male patients with transfusion-dependent beta-thalassemia. Pediatr Res 2018;83:655–61. https://doi.org/10.1038/pr.2017.296.

[15] Bhasin S, Cunningham GR, Hayes FJ, Matsumoto AM, Snyder PJ, Swerdloff RS, et al. Testosterone Therapy in Men with Androgen Deficiency Syndromes: An Endocrine Society Clinical Practice Guideline. J Clin Endocrinol Metab 2010;95:2536–59. https://doi.org/10.1210/jc.2009-2354.

[16] von Elm E, Altman DG, Egger M, Pocock SJ, Gøtzsche PC, Vandenbroucke JP. The Strengthening the Reporting of Observational Studies in Epidemiology (STROBE) statement: guidelines for reporting observational studies. Lancet 2007;370:1453–7. https://doi.org/10.1016/S0140-6736(07)61602-X.

[17] Chern JPS, Lin K, Tsai W, Wang S, Lu M, Lin D, et al. Hypogonadotropic Hypogonadism and Hematologic 2003;25:880–4.

[18] Chen MJ, Peng SSF, Lu MY, Yang YL, Jou ST, Chang HH, et al. Effect of iron overload on impaired fertility in male patients with transfusion-dependent beta-thalassemia. Pediatr Res 2018;83:655–61. https://doi.org/10.1038/pr.2017.296.

[19] Wu H, Lin C, Chang Y, Wu K, Lei R, Peng C, et al. Survival and complication rates in patients with thalassemia major in Taiwan. Pediatr Blood Cancer 2017;64:135–8.

[20] Daraghmeh NM. Management and Complications of Thalassemic Patients in Palestine: Retrospective Study 2016.

[21] Habeb AM, Al-Hawsawi ZM, Morsy MM, Al-Harbi AM, Osilan AS, Al-Magamsi MS, et al. Endocrinopathies in beta-thalassemia major. Prevalence, risk factors, and age at diagnosis in Northwest Saudi Arabia. Saudi Med J 2013;34:67–73.

[22] Ahmed E, Shaheen M. Prevalence of Hypogonadism in Thalassemia major patients in Gaza strip 2019.

[23] Belhoul KM, Bakir ML, Saned M-S, Kadhim AMA, Musallam KM, Taher AT. Serum ferritin levels and endocrinopathy in medically treated patients with β thalassemia major. Ann Hematol 2012;91:1107–14.

[24] Srisukh S, Ongphiphadhanakul B, Bunnag P. Hypogonadism in thalassemia major patients. J Clin Transl Endocrinol 2016;5:42–5. https://doi.org/10.1016/j.jcte.2016.08.001.

[25] Chatterjee R, Katz M, Cox TF, Porter JB. Prospective study of the hypothalamicLJpituitary axis in thalassaemic patients who developed secondary amenorrhoea. Clin Endocrinol (Oxf) 1993;39:287–96.

[26] Najafipour F. Evaluation of endocrine disorders in patients with thalassemia major 2008.

[27] Merchant RH, Shirodkar A, Ahmed J. Evaluation of growth, puberty and endocrine dysfunctions in relation to iron overload in multi transfused Indian thalassemia patients. Indian J Pediatr 2011;78:679– 83.

[28] Moayeri H, Oloomi Z. Prevalence of growth and puberty failure with respect to growth hormone and gonadotropins secretion in beta-thalassemia major 2006.

[29] Majeed MS. Evaluation of some Biochemical and Endocrine Profiles in transfusion-dependent Iraqi major β-thalassemia patients. Iraqi J Sci 2017;58:639–45.

[30] Safarinejad MR. Reproductive hormones and hypothalamic-pituitary-ovarian axis in female patients with homozygous β-thalassemia major. J Pediatr Hematol Oncol 2010;32:259–66.

